# Colorimetric Test for Fast Detection of SARS-CoV-2 in Nasal and Throat Swabs

**DOI:** 10.1101/2020.08.15.20175489

**Authors:** Bartolomeo Della Ventura, Michele Cennamo, Antonio Minopoli, Raffaele Campanile, Sergio Bolletti Censi, Daniela Terracciano, Giuseppe Portella, Raffaele Velotta

## Abstract

Mass testing is fundamental to face the pandemic caused by the coronavirus SARS-CoV-2 discovered at the end of 2019. To this aim, it is necessary to establish reliable, fast and cheap tools to detect viral particles in biological material so to identify the people capable to spread the infection. We demonstrate that a colorimetric biosensor based on gold nanoparticle (AuNP) interaction induced by SARS-CoV-2 lends itself as an outstanding tool for detecting viral particles in nasal and throat swabs. The extinction spectrum of a colloidal solution of multiple viral-target gold nanoparticles – AuNPs functionalized with antibodies targeting three surface proteins of SARS-CoV-2 (spike, envelope and membrane) – is redshifted in few minutes when mixed to a solution containing the viral particle. The optical density of the mixed solution measured at 560 nm was compared to the threshold cycle (*C_t_*) of a Real Time-PCR (gold standard for detecting the presence of viruses) finding that the colorimetric method is able to detect very low viral load with a detection limit approaching that of RT-PCR. Since the method is sensitive to the infecting viral particle rather than to its RNA, the achievements reported here open new perspective not only in the context of the current and possible future pandemics, but also in microbiology as the biosensor proves itself to be a powerful though simple tool for measuring the viral particle concentration.

---

Since its identification in China in late 2019, the SARS-CoV-2 epidemic has spread rapidly worldwide affecting millions of people, thus pushing the World Health Organization (WHO) to declare a COVID-19 outbreak a global health emergency. Mass testing is fundamental to identify and isolate clusters in order to limit and eventually eradicate SARS-CoV-2.^1^ The gold standard for diagnosing COVID-19 infection is a reverse transcription real-time polymerase chain reaction (RT-PCR)^2^ that is able to detect the virus genetic material (RNA) in samples collected via nasopharyngeal swab. Currently, only qualitative RT-PCR assays are available that yield positive/negative results without providing information about the viral load. Due to its complexity, RT-PCR tests are performed in certified laboratories, are time consuming, require experienced personnel, and can hardly lend themselves for mass screening.^3–6^ Huge efforts are put in overcoming such a bottleneck thereby making nucleic acid amplification suitable for Point Of Care tests, but the variety of methods^7^ and the lack of any commercial solution demonstrates that the gap between research and real applications is still to be filled.^8^ The main reason for that has to be found in the detection principle (RNA-extraction, reverse transcription and amplification) whose complexity, though greatly reduced by several approaches (e.g. Loop-mediated Isothermal Amplification (LAMP) and Recombinase Polymerase Amplification (RPA) or CRISPR-based detection) is far from being used for quick POC tests.

Lateral flow assays (LFA)s are among the actual biosensing platforms for home tests and potentially for mass screening,^9–12^ but the relatively poor sensitivity inherent to this technology^13^ makes urgent the quest for a different approach.^14^ Biosensors based on metal nanoparticles are often proposed because of their unique optical properties, which makes them potentially suitable to develop easy-to-use and rapid colorimetric diagnostic tests for point-of-care applications or even for home use.^15^ Due to its surface chemistry and given its biocompatibility gold is generally preferred to other metals.^16^ The physical process underlying this class of biosensors is the Localized Surface Plasmon Resonance (LSPR) that consists of coherent and non-propagating oscillations of free electrons in metal nanoparticles arising when they interact with an electromagnetic wave whose frequency resonates with the plasmonic one.^17,18^ Colorimetric detection based on gold nanoparticles (AuNPs) takes advantage of the color change occurring in a colloidal suspension from red to blue as a result of LSPR coupling among the nanoparticles.^19^ AuNP aggregation can be regulated using biological mechanisms such as antigen-antibody (Ab) interaction, in which case different strategies can be used to immobilize Abs correctly oriented on the surface of the AuNPs, although the complexity of the standard procedures makes them unsuitable for industrial applications on a large scale.^20^

The Photochemical Immobilization Technique (PIT) is a surface functionalization procedure that only requires UV activation of the Abs and leads to a high density functionalized surface within minutes.^21,22^ PIT has proven itself to be effective in tethering Abs upright not only on flat surfaces,^23–26^ but also on AuNPs, which were used either to ballast small antigen^27^ or to realize a colorimetric biosensor for detecting IgGs^28^ and estradiol.^29^ In the latter cases, the presence of the antigen was detected as a change in the absorbance that can be easily measured by a spectrophotometer or even by naked eye. An approach relying on nanoparticle aggregation induced by the presence of the antigen was also used to detect the influenza A virus, but no clinical application was reported to demonstrate the effectiveness of the whole procedure in clinical cases.^30^

Here, we report on the realization of a colorimetric biosensor that can be used for COVID-19 mass testing with sensitivity and specificity higher that 95% as demonstrated by a comparative analysis carried out on a total of 94 samples (45 positive and 49 negative), tested by standard RT-PCR in the Virology Unit of A.O.U. Federico II/Department of Translational Medicine of the University of Naples “Federico II”. The detection scheme is shown in Figure 1a and consists of a colloidal solution of PIT-functionalized AuNPs (f-AuNPs) against three surface proteins of SARS-CoV-2: spike, envelope and membrane (S, E, and M, respectively, in Figure 1a). AuNP fabrication (20 nm diameter), characterization and functionalization are described in the Sup porting Information (see sections S1-S5), that contains a scalable procedure to realize the colloidal solution for COVID-19 test. In this approach, the sample was a solution of Universal Transport Medium (UTM^®^, Copan Brescia, Italy), in which the specimen was dipped after its collection from the patient and without any additional treatment (see Supporting Information S6). Our test was carried out by mixing 50 μL of the f-AuNP colloidal solution with 100 μL of sample and 100 μL of ultrapure water. The presence of the viral particles (virions) induced the formation of a nanoparticle layer on its surface (Figure 1b) that led to a redshift of the optical density (OD) in the extinction spectrum of the solution (Figure 1c). When the viral load was relatively high, i.e. *C_t_* < 15 (Supporting Information S7), the color change from red to purple was visible even by naked eye (Figure 1a-b).

**Figure 1.**
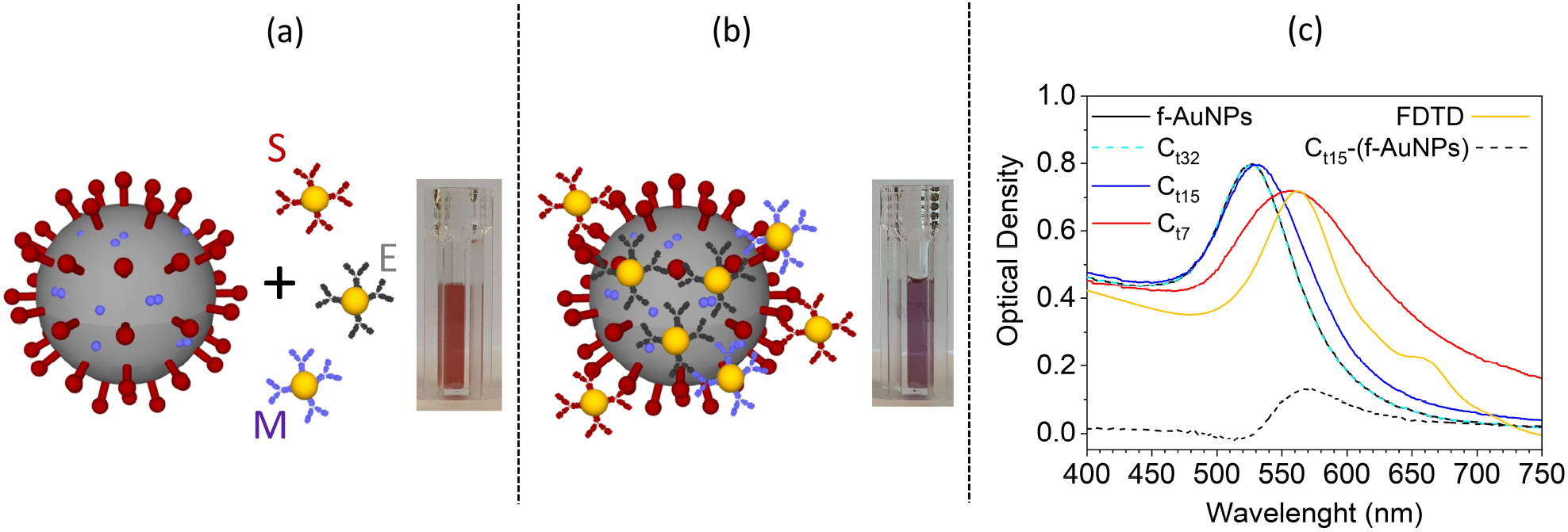
(a) Sketch of the SARS-CoV-2 and functionalized AuNPs. SARS-CoV-2 proteins (spike, membrane and envelope) and their corresponding antibody (S, E, and M) are highlighted in dark red, light violet and gray, respectively. The inset shows the pink colloidal solution containing the anti-SARS-CoV-2 functionalized AuNPs (f-AuNPs). (b) The f-AuNPs surround the virion forming a nanoparticle layer on its surface. Their interaction leads to a shift of the resonance peak in the extinction spectrum and, hence, to a color change visible in the inset. (c) Extinction spectra reporting the OD of f-AuNP colloidal solution mixed with samples from patients with different viral load. At very low virion concentration (curve C_t32_) the extinction spectrum is not distinguishable from the spectrum of f-AuNPs (black continuous line). At intermediate virion concentration (curve C_t15_) the extinction spectrum is slightly red-shifted and its difference with the “control” (f-AuNPs) produces the curve C_t15_-(f-AuNPs) that evidences the contribution entailed by the virion. At high virion concentration (curve C_t7_), the extinction spectrum peaks at 560 nm as for C_t15_-(f-AuNPs). The agreement between the curve C_7_ and the simulated spectrum (gold continuous line, scaled to the experimental one) from a dielectric sphere (100 nm diameter) surrounded by smaller AuNPs (20 nm diameter) confirms the interpretation of the extinction spectra as due to nanoparticle aggregation.

The extinction spectrum of f-AuNPs reported in Figure 1c (black continuous line) is not distinguishable from the spectrum of a mixed solution with a sample having *C_t_* = 32 (dashed light blue curve, C_t32_). On the contrary, red shift is observed for a sample with *C_t_* = 15 (blue continuous line, C_t15_), which becomes much more noticeable when *C_t_* = 7 (red continuous line, C_t7_). The contribution of the virion (surrounded by nanoparticles) to the extinction spectrum can be deduced by subtracting the spectrum of f-AuNPs from C_t15_. In fact, the curve C_t15_-(f-AuNPs) shows a peak at a wavelength comparable to that exhibited by C_t7_ (approximately 560 nm).

To further confirm that the spectrum C_t7_ arose from f-AuNPs surrounding SARS-CoV-2, we simulated the virion as a 100 nm diameter^31,32^dielectric sphere of 1.45 refractive index.^33^ A change able number of gold nanoparticles (20 nm diameter)^34^ was randomly positioned on the sphere (Supporting Information S8). We implemented the “FDTD solutions” tool in Lumerical software that provides numerical solutions of the Maxwell’s equations by finite-difference time-domain (FDTD) method within a Mie problem-like workspace. A sketch of the simulation workspace is depicted in Figure S8.1a, whereas Figure S8.1b shows the LSPR wavelength as a function of the number of AuNPs on the virion surface. The plasmon resonance redshifts as the number N of AuNPs onto the virion surface increases (Figure S8.1b). The maximum number of AuNPs that can be placed on a perfectly spherical surface with 100 nm diameter is N = 80, but the steric hindrance offered by surface proteins like the spike one surely limits the filling capacity of the virion surface. In fact, from Figure S8.1b we see that the experimental value of *λ*_LSPR_ = 560 nm is achieved when *N* = 70, a value only slightly smaller than the maximum achievable. The extinction spectrum corresponding to *N* = 70 AuNPs (scaled to the experimental one) is reported in Figure 1c (golden continuous line) and shows a more than satisfactory agreement with the experimental one (red continuous line), thereby confirming that the simple model proposed here is able to capture the essential physical processes underlying the virion detection.

In order to test the validity of the colorimetric biosensor, we analyzed real samples previously examined by RT-PCR. The samples were from 45 positive to SARS-CoV-2 patients for which *C_t_*≤35 and 49 negative patients (*C*_t_>35). For all of them, we measured the optical density at 560 nm (OD_560_) by a commercial microplate reader (Figure 2a). It is quite evident the correlation between the *C_t_* value (reported on the top scale) of the positives (red circles) and OD_560_, whereas all the negatives (identified by a progressive number in the bottom scale) are randomly distributed providing a “control” value of 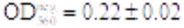. Also shown in Figure 1a is a horizontal line at an OD_560_ value of 0.252, which makes clear the ability to discriminate positives from negatives offered by our colorimetric biosensor. In fact, with such a threshold we get 96% and 98% for sensitivity and specificity, respectively. This result is even more remarkable if we consider that *C_t_*>30 correspond to a very low viral load for which the infection aptitude is questioned. To this aim, it is important noticing that in the early phase of the infection, high viral loads are detected by RT-PCR in upper respiratory specimens with low *C_t_* readouts (*C_t_*< 20, with our assay). In the late phase of the infection, a dramatic drop in viral loads is observed with higher *C_t_* readouts (*C_t_*>30 with our assay). Positive results with high *C_t_* readouts pose a diagnostic challenge, since they do not necessarily indicate active infection by a replicating virus. It has been observed using viral culture that patients with high *C_t_* RT-PCR and protracted positivity are not infectious, suggesting that the assay likely detects non active viral particles such as genetic material present in remnants of inactive virus^35^ thereby making our approach of high diagnostic value.

**Figure 2.**
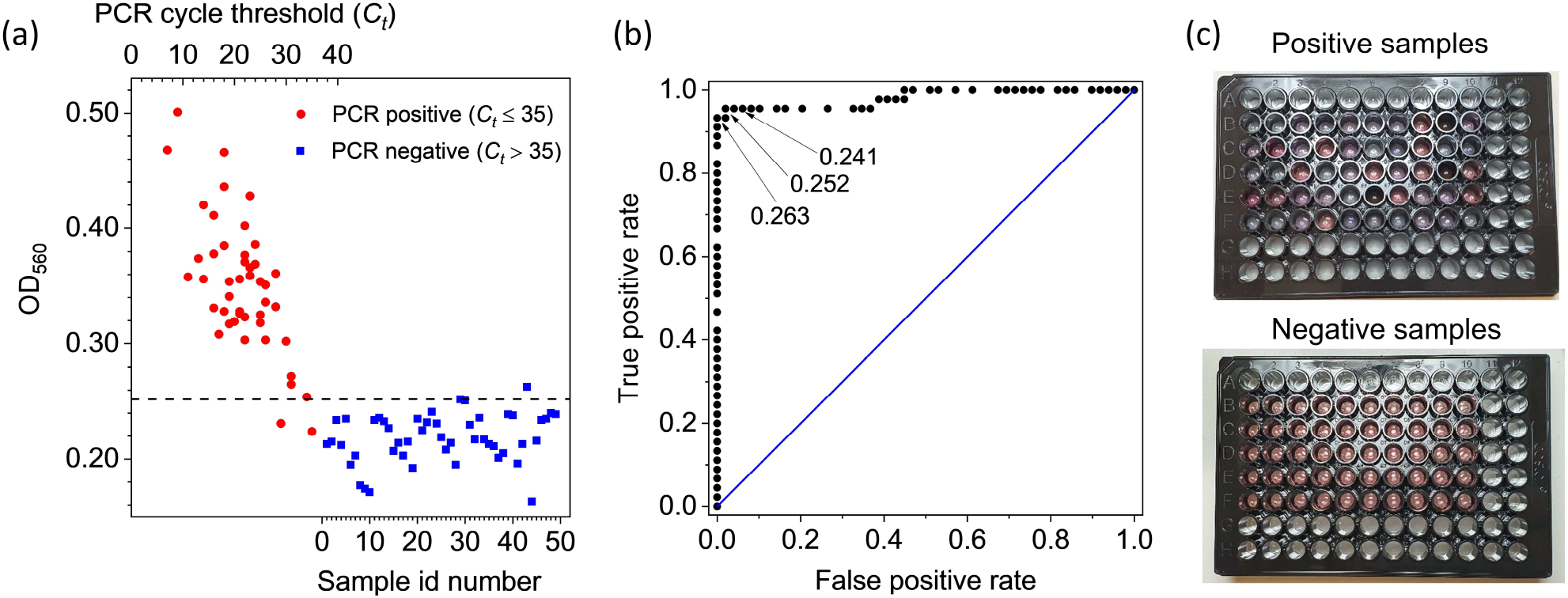
(a) Results of the colorimetric test on real thawed samples from 45 positive (red circle points) and 49 negative patients (blue square points) previously tested by RT-PCR. For all of them the extinction coefficient was measured at 560 nm. The positive samples (C_t_ ≤ 35) are identified in the plot by their RT-PCR cycle threshold (top scale), whereas the negative sample (C_t_ > 35) are simply numbered (bottom scale). The horizontal line at 0.252 extinction coefficient would lead to a test with 96% sensitivity and 98% specificity. (b) ROC curve retrieved from the data of the panel (a). The area under the curve is 0.98. Also shown three threshold values for the extinction coefficient that would provide the following sensitivity and specificity: 96% and 94% (0.241), 96% and 98% (0.252), and 94% and 100% (0.263), respectively. (c) Picture of the 96 multiwell plate containing 250 μL of positive (top panel) and negative (bottom panel) samples. The plate reading was carried out by a commercial multiwell reader that took less than 1 min.

The Receiver Operating Characteristic (ROC) curve from the data shown in Figure 2a is reported in Figure 2b together with the indication of three threshold values, all of them leading to sensitivity and specificity significantly higher than 90%. In particular, the highest threshold value (0.263) leads to 100% specificity while keeping the sensitivity at the remarkable value 94%. Overall, the high performance of the test associated to the colorimetric biosensor is demonstrated by the area under the ROC curve whose value is 0.98. The qualitative difference in the color between positives and negatives can be observed in Figure 2c that shows a picture of the multiwells containing the samples whose analysis is summarized in Figure 2a-b.

To measure the dose-response curve of the biosensor, we assessed the optical density 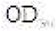 of samples obtained by serial dilutions (1:10) of an initial volume with very high viral load (*C_t_* = 7). The results are shown in Figure 3, in which 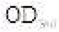 is reported as a function of the relative concentration of SARS-CoV-2. For convenience, the top scale reports the equivalent *C_t_* obtained by considering that 1:10 dilution corresponds to 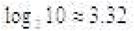 change in *C_t_*, whereas the vertical dashed blue line identifies the starting concentration. As it turns out, a detectable signal can be appreciated even after 7 serial dilutions (1:10^7^) confirming the wide detection range resulting from the validation measurements reported inFigure 1. In particular, from Figure 3 we can safely assess that our limit of detection lies above *C_t_* = 30 thereby corroborating the reliability of the measurements reported in Figure 2a even when they refer to high *C_t_*.

**Figure 3.**
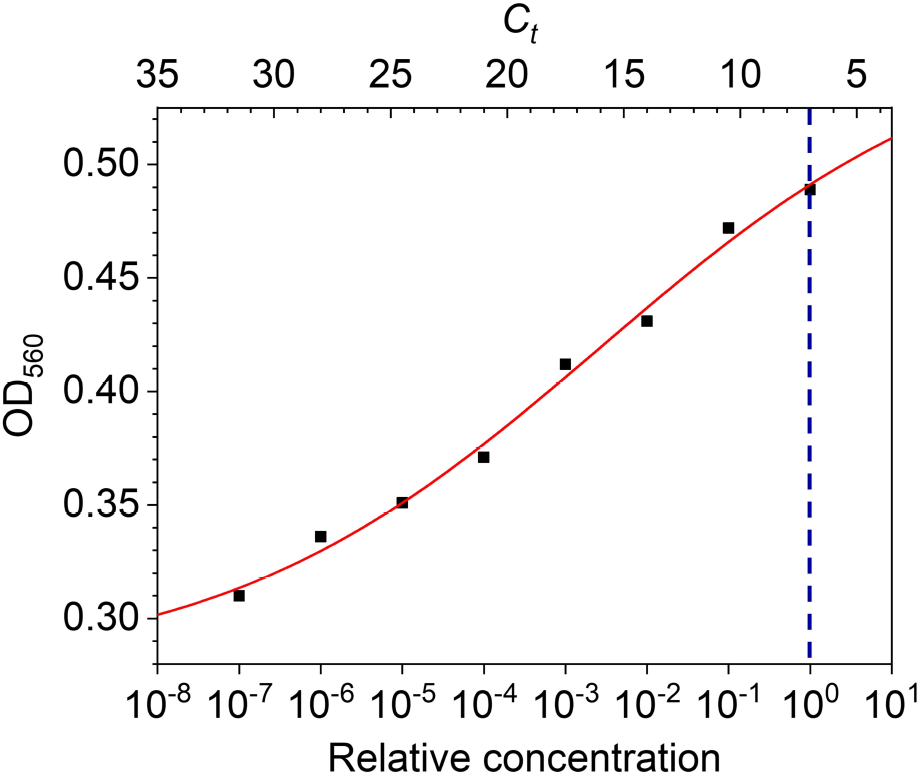
Optical density of the solution measured at 560 nm as a function of SARS-CoV-2 concentration (bottom axis). The virion concentration was obtained by serial dilution starting from a threshold cycle value *C_t_* = 7 (vertical dashed blue line). Each decrease by a decade in virion concentration (bottom axis) corresponds to an increase of the nominal *C_t_* of approximately 3.32 (top axis). The red continuous line is the best fit of the experimental data by Hill equation. The uncertainty in the reading is taken as the resolution of the instrument and the error bar is within the data point.

The behavior of the 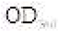 as a function of the virion concentration C reported in Figure 3 can be described by the four parameters Hill equation^36^

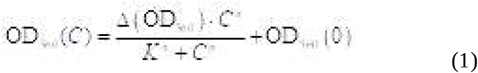

In Equation 1, 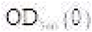 is the OD of the “control”, 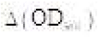 is the maximum OD_560_ variation, *K* is the concentration (in this case it is a relative concentration) at which the OD_560_ variation reaches 50% of its maximum and *n* is the so-called Hill’s coefficient.^37^ The best fit of the experimental data with Equation 1 yielded the curve in Figure 3 and the following values for the parameters: 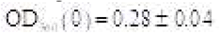, 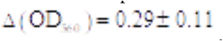, 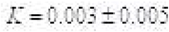 and 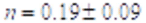.

The (relative) concentration *K* is essentially undetermined as a consequence of the high detection range of our method, which entails small response to the concentration changes (low sensitivity), and hence, high uncertainty on the concentration measurements.

A value for *n* significantly smaller than 1 indicates the occurrence of a negatively cooperative binding, a behavior which we expect by considering that the probability a f-AuNP binds the virion reduces as the surface covering grows. While 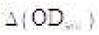 simply provides information about the asymptotic value of the optical density, 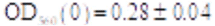 is an independent measurement of the OD when no virions are present. It turns out that 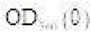 is fully compatible with 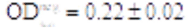 previously measured thereby confirming the consistency of our description.

In conclusion, we realized a colorimetric biosensor based on a colloidal solution of AuNPs (20 nm, OD≈1) each of them functionalized with Abs against one of the three surface proteins of SARS-CoV-2 (spike, envelop and membrane). The ratio among the three kinds of functionalized AuNPs was 1:1:1. Although both the ratio and the size of AuNPs are still susceptible to optimization so to allow one to push even further the limit of detection, the current performances of the biosensor would already permit its use as a test for mass screening since the detection is based on the interaction among the virions and the pAb-functionalized AuNPs (single step detection) without any pretreatment (e.g. RNA extraction and amplification). The comparison of the readout of our biosensor at 560 nm with the threshold cycle (*C_t_*) of a RT-PCR proved that viral loads corresponding to *C_t_* = 30 are detected by the colorimetric biosensor. This threshold is of particular importance because it corresponds a low viral load for which the infecting capacity is likely negligible.^35^ Such a good performance has to be ascribed to a high filling ratio of the virion surface that results from the presence of multiple Abs (three proteins are targeted) and an effective AuNP surface functionalization procedure (PIT). In fact, through PIT not only one Fab is always exposed so to make AuNP highly “reactive”, but also the Abs are attached to the surface (side-on position) without any linker (e.g. protein A), the latter being detrimental for the plasmonic interactions among AuNPs on which the colorimetric biosensor is based.

Another remarkable feature of the biosensor described here relies on its sensitivity to the virion rather than to its content (RNA). The importance of this is twofold: 1) after the calibration of the optical response, the biosensor lends itself as a powerful tool to quantify the viral load, a non-trivial issue in diagnostic assays in virology; 2) being sensitive only to the virions, the biosensor detects the presence of active viral particles; thus, our method is apt to assess the actual degree of infectiveness of a specimen. Conversely the qualitative RT-PCR assays do not allow to clearly discriminate between high infective samples from those containing inactive virus.

As a final remark, we point out that the colorimetric solution described here can be easily modified to target other viruses. Thus, we expect that single-step colorimetric detection of viruses can become a general technique to be used for laboratory applications as well as point-of-care testing.

## Data Availability

The data are available upon request to the corresponding authors

## ASSOCIATED CONTENT

### Supporting Information

Materials (S1). Instruments (S2). Gold nanoparticle synthesis (S3). Optical and morphological AuNP characterization (S4). Functionalization (S5). Storage of the samples and validation of the measurements with PCR (S6). Threshold cycle in PCR measurement and viral load (S7). FDTD optical simulations (S8).

This material is available free of charge via the Internet at http://pubs.acs.org.

## AUTHOR INFORMATION

### Notes

The authors declare no competing financial interests.

## ACKNOWLEDGMENT

We thank dr Luigi Atripaldi, dr Roberto Parrella and dr Claudia Tiberio for their availability to carry out preliminary tests in the Laboratory of Microbiology at Hospital “Domenico Cotugno” in Naples.

## Notes

### Competing Interest Statement

The authors have declared no competing interest.

### Funding Statement

The consumables were partly purchased by Cosvitec scarl

### Author Declarations

This study was approved by the IRB of the University of Naples Federico II with the document n. 140/20.

